# Bicuspid valve CT Registry of balloon expandable TAVR: BETTER TAVR Registry

**DOI:** 10.1101/2023.07.31.23293458

**Authors:** Jorge Chavarria, Felipe Falcao, Hatim AlRaddadi, Amir Aziz, Alexander Dick, Kevin Chung, Janar Sathananthan, David Meier, Noman Ali, James Velianou, Madhu Natarajan, Iqbal Jaffer, David Wood, Neil Fam, Tej Sheth

## Abstract

**Objectives:** We evaluated determinants of stent geometry in bicuspid valves treated with balloon expandable TAVR.

**Background:** The anatomic substrate of bicuspid valves may lead to suboptimal TAVR stent expansion and geometry.

**Methods:** A multicentre observational registry of patients who underwent post TAVR CT to determine stent area (vs nominal valve area) and stent ellipticity (maximum diameter/minimum diameter). Predictors of relative stent expansion (minimum area/average of inflow + outflow area) and stent ellipticity were evaluated in a multivariable regression model including valve calcium volume, presence of raphe calcium, sinus diameters indexed for area derived annular diameter, and performance of pre-dilation and post-dilation.

**Results:** The registry enrolled 101 patients from 4 centres. Minimum stent area (vs nominal area) was 88.1% and maximum ellipticity was 1.10 with both observed near the midframe of the valve in all cases. Relative stent expansion ≥90% was observed in 64/101 patients. The only significant predictor of relative stent expansion ≥90% was performance of post-dilation (p<0.01). Relative stent expansion ≥90% was seen in 86% of patients with post dilation compared to 57% of without (p<0.001). The stent ellipticity ≥ 1.1 was observed in 47/101 patients. The significant predictors of stent ellipticity ≥1.1 was the presence of raphe calcification (p=0.02) and intercommisural diameter at 4 mm above the annular plane (p=0.004).

**Conclusion:** Relative stent expansion ≥90% was associated with the performance of post-dilation and stent ellipticity was associated with the presence of calcified raphe and sinus dimensions (Intercommisural diameter at 4 mm above the annular plane). Further studies to determine optimal deployment strategies in bicuspid valves are needed.

Graphical abstract
Predictors of stent expansion and ellipticity from a multicenter registry of patients with bicuspid anatomy undergoing TAVR with balloon expandable valves.

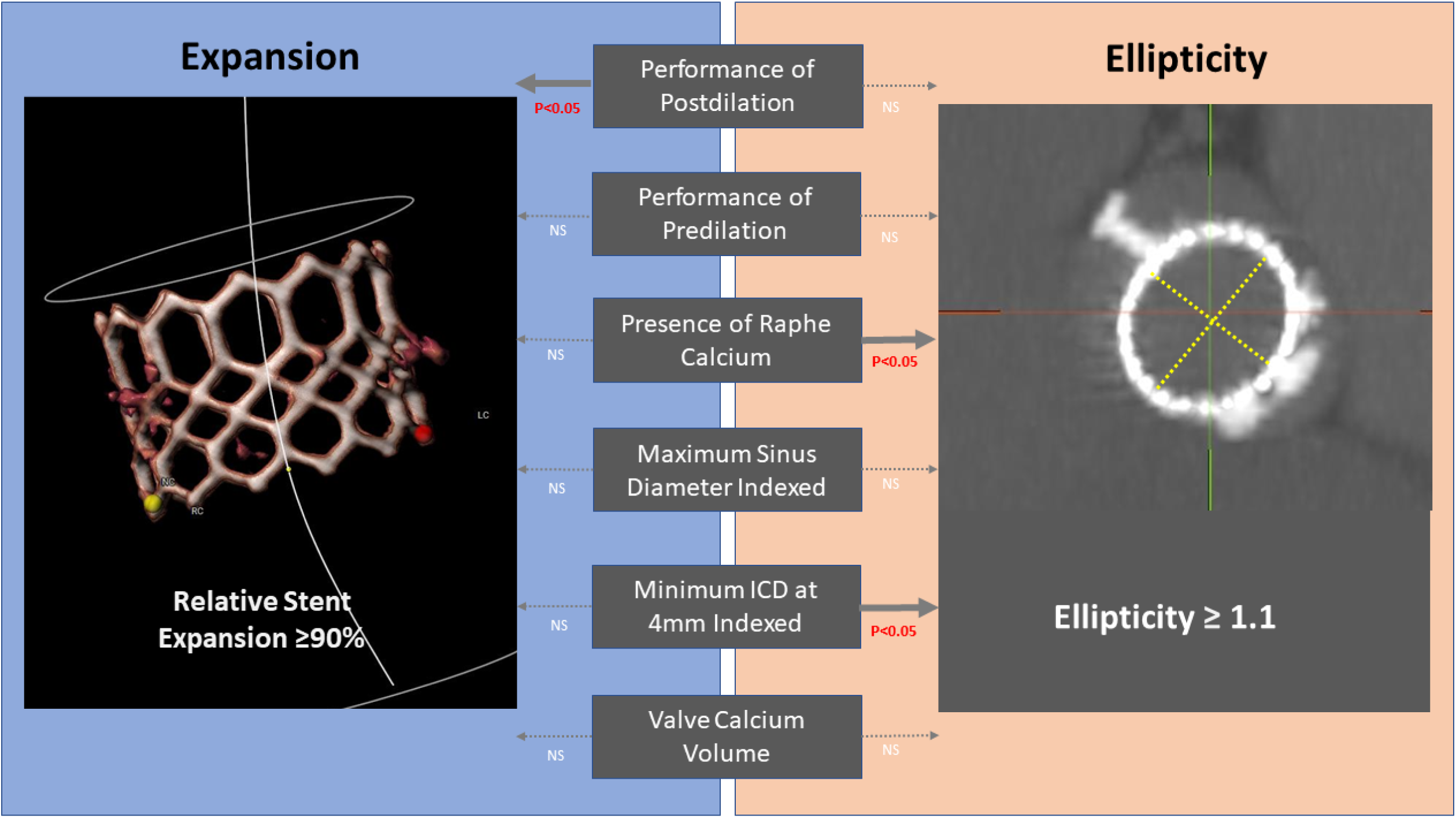

## Background

Bicuspid Aortic Valves (BAV) are the most common congenital valvular abnormality, with a prevalence estimated between 0.5 to 2%^1,2^. BAV account for 50% of patients referred for Aortic Valve Replacement, and 22% to 28% of the patients above 80 years of age referred for Surgical Aortic Valve Replacement (SAVR)before the TAVR era^3^. BAV are more commonly encountered in the treatment of younger patients at low surgical risk. In the TVT registry, balloon expandable TAVR of bicuspid valves accounted for <5% of all TAVRs performed in low-risk patients^4^. In this selected population, procedural complications were extremely low and comparable to tricuspid valve patients. Hemodynamic results including gradients and paravalvular leak were also similar in both valve types. Although these acute procedural outcomes are excellent, the unique anatomic substrate of bicuspid valves has raised concern about potential suboptimal stent geometry and its impact on valve hemodynamics, durability, and repeatability. Stent under-expansion may be associated with hypoattenuation leaflet thickening and worse outcomes in the long-term^5,6^. Several CT follow-up studies have shown that BEV have more stent ellipticity and lower stent expansion compared to tricuspid valves^2,7^ while another showed similar ellipticity, expansion, and foreshortening as tricuspid valves ^8^.

Determinants of stent geometry in bicuspid TAVR performed with BEV have not been well studied. It is possible that leaflet and raphe calcification may impact on stent geometry^7^ and that post-dilation may help to overcome the resistance posed by the bicuspid leaflets and improve stent expansion and symmetry. If optimal stent expansion can be predicted from pre-TAVR CT images or achieved through procedural techniques, it may help to ensure more predictable results in the treatment of bicuspid valve patients with TAVR. We, therefore, evaluated the determinants of stent expansion and ellipticity in patients with bicuspid valves treated with balloon expandable TAVR using pre and post TAVR CT images.

## Methods

We included patients undergoing balloon expandable TAVR with a Sapien 3 or Sapien 3 Ultra valve in the setting of severe aortic stenosis in bicuspid valve disease with available follow-up CT imaging from four high volume academic Canadian TAVR centres. The selection of bicuspid cases for TAVR treatment and follow-up CT imaging was at the discretion of each site’s heart team. All included patients underwent post TAVR CT imaging to assess valve size and geometry. Patients undergoing post-TAVR CT imaging specifically to evaluate for leaflet thrombosis due to elevated postoperative gradients were excluded. All patients underwent pre-TAVR CT evaluation for routine pre-procedure planning. The study was approved by the research ethics board.

### Post TAVR CT Evaluation

Image analysis was performed with 3Mensio (Pie Medical, Netherlands) review workstation and is illustrated in Figure 1. CT analysis performed by an experienced CT reader. Measurements were taken in the cardiac phase with best image quality. A manual 3D tool of a dedicated software for TAVR analysis was used (3mensio Medical Imaging BV, Netherlands). Two orthogonal viewing planes were aligned with the long axis of the valve stent to obtain a short axis image in the third viewing plane. The stent frame was assessed at 4 cross-sectional levels (inflow, minimal expansion level, mid-stent and outflow). To ensure consistency in area measurement regardless of the extent of blooming artifact interference, the stent area was traced manually by connecting points placed in the center of each stent strut. An area derived diameter was determined from this initial area estimate and increased by 0.6mm to account for strut thickness. It was then converted back to the final valve area estimate. Relative stent expansion was defined as the CT-derived minimal area divided by the average of the outflow and inflow areas 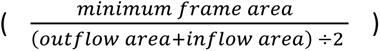. Ellipticity was calculated as the maximum stent diameter/minimum stent diameter at 4 cross-sectional levels (inflow, minimal expansion level, mid-stent and outflow). The largest ratio of ellipticity was used for multivariate analysis. The presence of hypoattenuation leaflet thickening was also noted.

**Figure 1:**
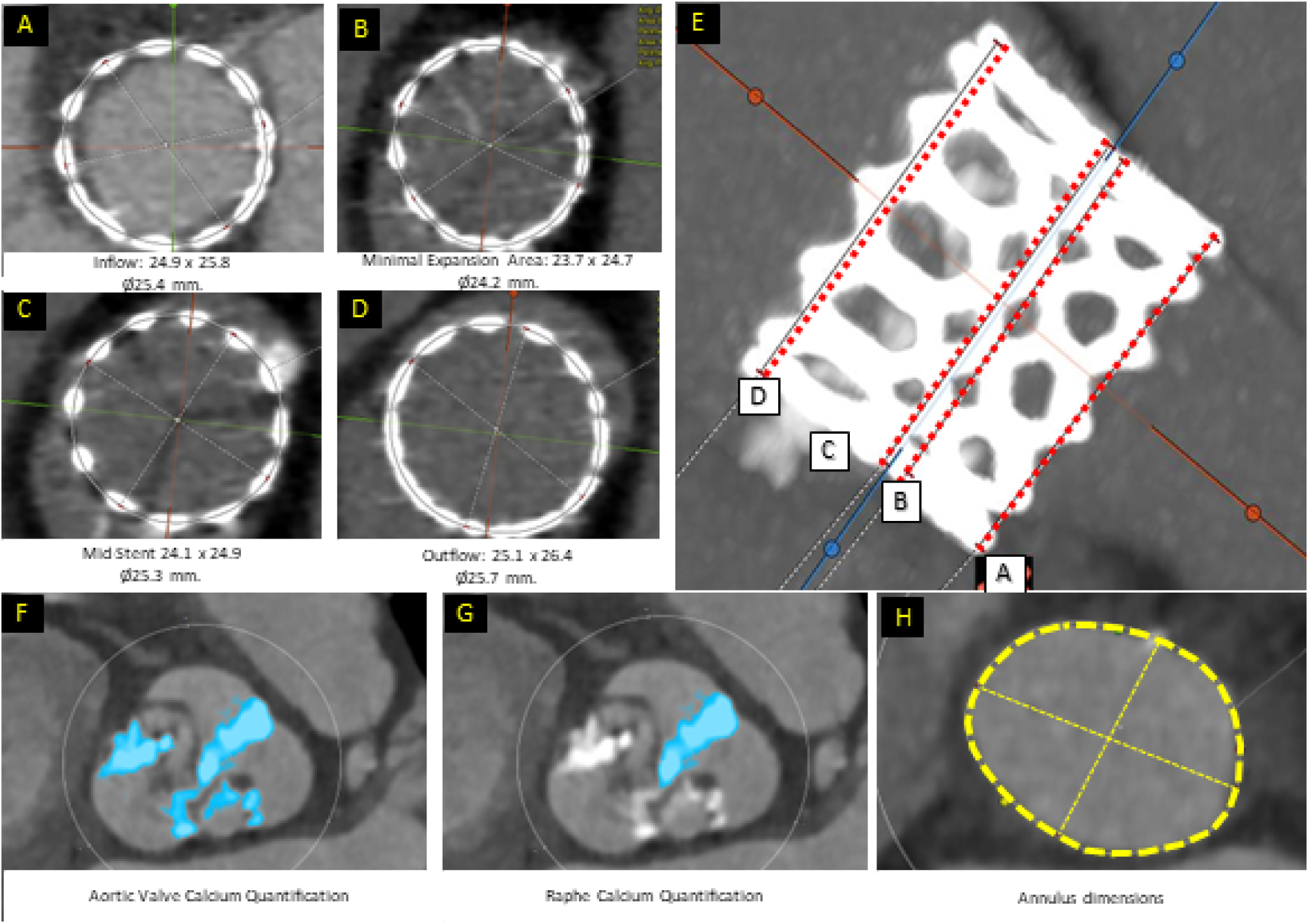
Definitions of post stent and pre-stent measures. Images show stent expansion at 4 levels. A: inflow, B: minimal expansion level, C: midframe D: outflow. The stent area was traced manually by connecting points placed in the center of each strut. E: Three-dimensional visualization of the Sapien valve. F: Total leaflet calcium volume, G: Raphe calcium volume, H: The annulus dimensions measures with minimum and maximum diameter.

### Pre TAVR CT Evaluation

The type of bicuspid valve was determined using the Sievers classification from the contrast CT images. Sievers 1 bicuspid were further classified by the location of the raphe. To quantify calcium volume in the contrast images, a dynamically adjusted HU unit threshold was chosen that was adjusted to 2 standard deviations above the contrast density in the aorta^9^. This threshold was then applied to quantify calcium volume for the valve leaflets. The raphe was described as calcified or noncalcified. The minimum and maximum sinus diameter was measured at the leaflet level. Annular area derived diameter was determined using standard techniques and divided into the sinus diameters to produce an indexed value for sinus dimensions. Intercommissural distance (ICD) were assessed 4 mm above the aortic annulus. The left ventricular outflow tract area was measured2 mm below the virtual annular plane.

### TAVR Procedure

TAVR was usually performed with conscious sedation and local anesthesia. Both pre and post-dilation were at the discretion of the operator.

### Follow-up Echocardiography

Data from the first post TAVR echocardiogram were included. All echocardiograms were interpreted by level 3 echocardiographers. The severity of PVL was graded as none, trace, mild, moderate, and severe according to Valve Academic Research Consortium 3 criteria^10^. Interpretation was performed by expert readers blinded to valve sizing parameters and angiographic results.

### Statistical Analyses

Baseline demographics were summarized using descriptive statistics. Continuous variables were expressed using means and standard deviations, and categorical variables using counts and percentages. Correlation was assessed with Pearson coefficients. Univariate and multivariate analysis were performed to evaluate pre-TAVR and procedural variables to predict relative stent expansion and stent ellipticity. Predictors of interest were total calcium burden, presence or absence of raphe calcium, maximum sinus diameter (indexed to area derived annular diameter), minimum ICD at 4 mm diameter (indexed to area derived annular diameter), performance of pre-dilation andpost-dilation. We assessed for collinearity between predictive variables using the variance inflation factor (VIF) and Tolerance statistics. If VIF is >10, we excluded one of the highly correlated variables. Potential predictors listed above will be forced in a multilinear regression model. With 101 patients, in order to detect an effect with an f^2^ = 0.15 at a power of (1-β) = 0.87 and type 1 error of 0.05, a critical F value was calculated at 2.4 We will then carry out forward stepwise generation of a logistic regression model by sequentially testing each hypothesized variable at the p<0.15 level and reassessing each previously included variable for eligibility to remain in the model at the p<0.1 level following each new variable assessment. For all analyses, statistical significance was considered when the p-value was less than 0.05. Statistical analyses were performed using STATA version 15.0 (StataCorp LP, College Station, Texas).

## Results

We evaluated 101 treated bicuspid patients who underwent TAVR with a BEV and had a follow-up CT performed. The baseline characteristics are shown in table 1. Most patients had Sievers 1 bicuspid valves with left right fusion. Procedural characteristics are shown in table 2. All most all procedures were performed with percutaneous femoral access and conscious sedation. Post-dilation was performed in 21% of patients, mostly with the same balloon volume.

**Table 1.** Baseline characteristics of patients with bicuspid aortic valve undergoing TAVR and CT follow-up imaging.

| Baseline characteristics | N= 101 |
| --- | --- |
| Age (SD) | 74.9 (7.6) |
| Male, gender (%) | 51 (50.5) |
| STS Score (%) | 2.1 (1.3) |
| Bicuspid Valve Type | 101 (100%) |
| Sievers 0 | 13 (12.9) |
| Sievers 1 with Left-Right Cusp Fusion | 64 (63.4) |
| Sievers 1 with Right-Non Cusp fusion | 24 (23.8) |
| Sievers 1 With Non-Left Cusp fusion | 0 (0) |
| BMI, (SD) | 1.9 (0.2) |
| Prior Revascularization (%) | 13 (12.9) |
| Prior Pacemaker (%) | 7 (6.9) |
| COPD (%) | 14 (13.9) |
| Prior Stroke | 14 (13.9) |
| GFR < 30 ml/min. | 0 (0) |
| DM (%) | 16 (15.8) |
| HTN (%) | 73 (72.3) |
| Dyslipidemia (%) | 62 (61.4) |
| Atrial Fibrillation (%) | 20 (19.8) |
| Mean Gradient, mmHg (SD) | 47.0 (14.8) |
| AVA, cm2 (SD) | 0.8 (0.2) |
| Ejection fraction, % (SD) | 57.7 (9.5) |
| Annular Area, mm2 (SD) | 527.1 (104.1) |
| LVOT Area, mm2 (SD) | 520.4 (112.9) |
| ICD at 4 mm above the annulus, mm | 28.0 (3.0) |
| ICD at 8 mm above the annulus, mm | 28.9 (2.9) |
| Ascending Aorta, maximum diameter above STJ, mm | 38.1 (5.1) |
| Aortic valve non contrast calcium score, Agatston (SD) | 2394.1 (1625.5) |
| Aortic valve density, AU/mm2, (SD) | 641.8 (325.7) |
| Aortic valve contrast volume, mm3 (SD) | 1043.6 (633.8) |

AU: Agatston units, SD: standard deviation, GFR : glomerular filtration rate, BSA: Body surface area,
AVA: Aortic Valve Area. LVOT: Left Ventricle Outflow Tract. ICD: Intercomissural diameter. STJ:
Sinotubular junction

**Table 2.**
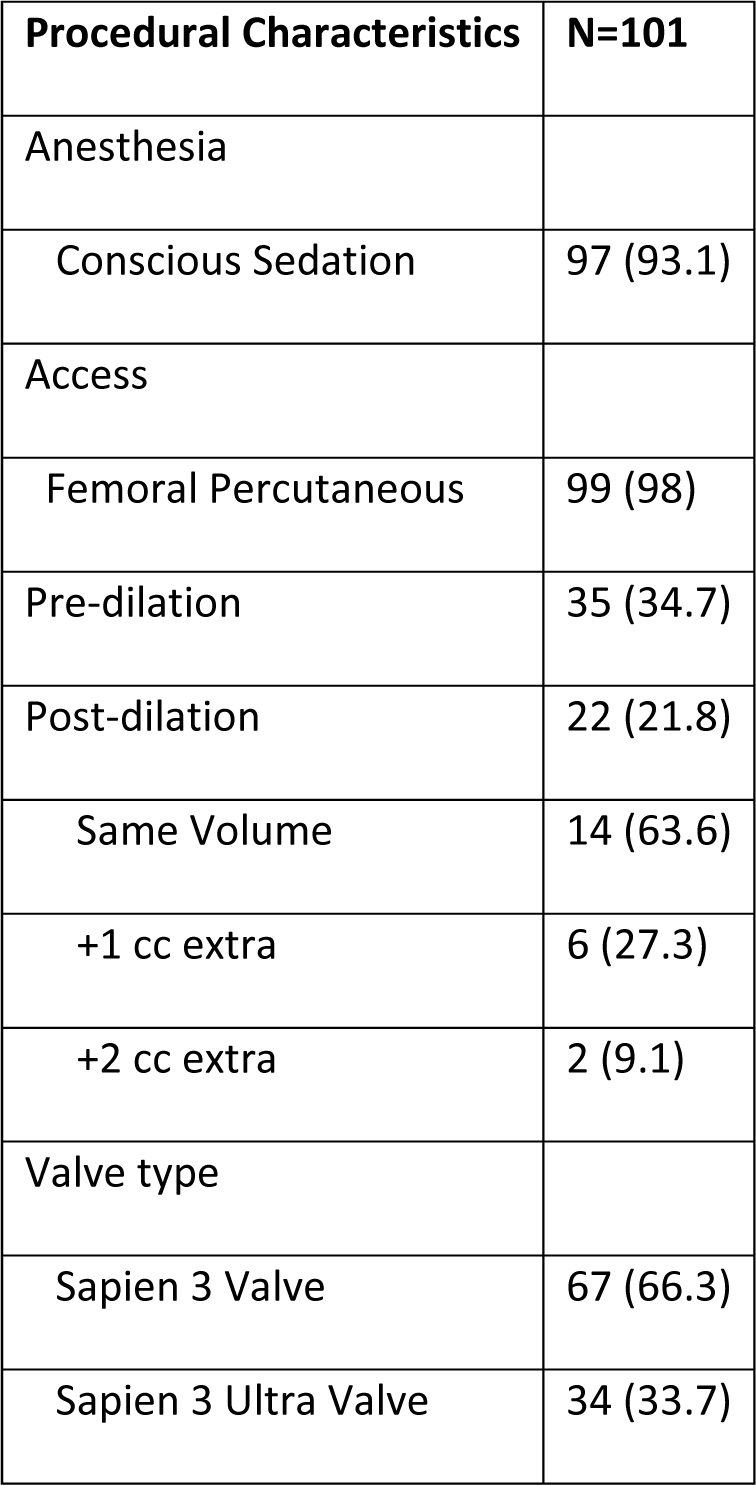
Procedural Characteristics.

| **Procedural Characteristics** | **N=101** |
| --- | --- |
| Anesthesia |  |
| Conscious Sedation | 97 (93.1) |
| Access |  |
| Femoral Percutaneous | 99 (98) |
| Pre-dilation | 35 (34.7) |
| Post-dilation | 22 (21.8) |
| Same Volume | 14 (63.6) |
| +1 cc extra | 6 (27.3) |
| +2 cc extra | 2 (9.1) |
| Valve type |  |
| Sapien 3 Valve | 67 (66.3) |
| Sapien 3 Ultra Valve | 34 (33.7) |

### Stent Geometry

Stent expansion as a percentage of nominal area was 95.4% at the inflow, 88.1% at the minimum area, 88.4% at the midframe and 99.7% at the outflow. Stent ellipticity was 1.07 at the inflow, 1.10 at the minimum area, 1.09 at the mid frame, and 1.07 at the outflow (Figure 2). The minimum stent area and maximum ellipticity were seen near the midframe of the valve in all cases.

**Figure 2.**
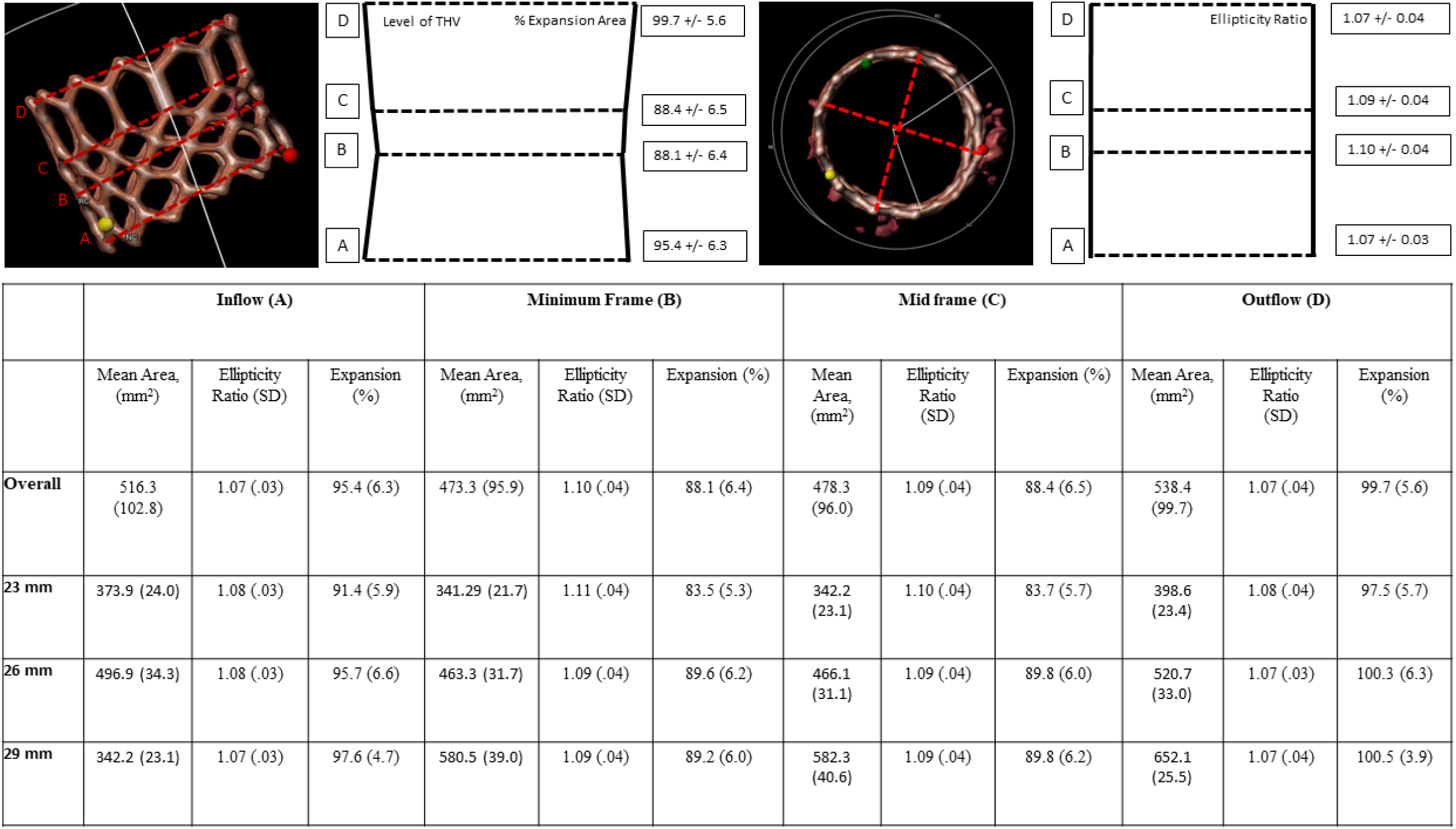
Measures of Valve Stent Geometry. Stent expansion relative to nominal stent area overall and by valve size (23, 26, and 29mm) at the inflow, midframe, minimum area, and outflow. Stent ellipticity at the inflow, midframe, minimum stent area, and outflow. Mean values and standard deviations (SD) are provided in the table.

### Predictors of Stent Expansion

Relative stent expansion ≥90% was achieved in 64/101 patients and mean relative stent expansion was 90.53% +/-3.21. Results of the multivariate model to identify factors associated with relative stent expansion are shown in Table 3. The only significant predictor of relative stent expansion was the performance of post-dilation. In contrast, there was no association with sinus dimensions, leaflet calcium volume, performance of pre-dilation, or the presence of raphe calcification (Figure 3). Among patients undergoing post-dilation, ≥90% relative stent expansion was observed in 86% of patients compared to 57% of patients who did not undergo post-dilation (p<0.001) (Figure 4). The interaction of stent expansion and post operative gradients is shown in Figure 5. There was weak negative correlation between expansion and mean gradients that was statistically significant

**Figure 3:**
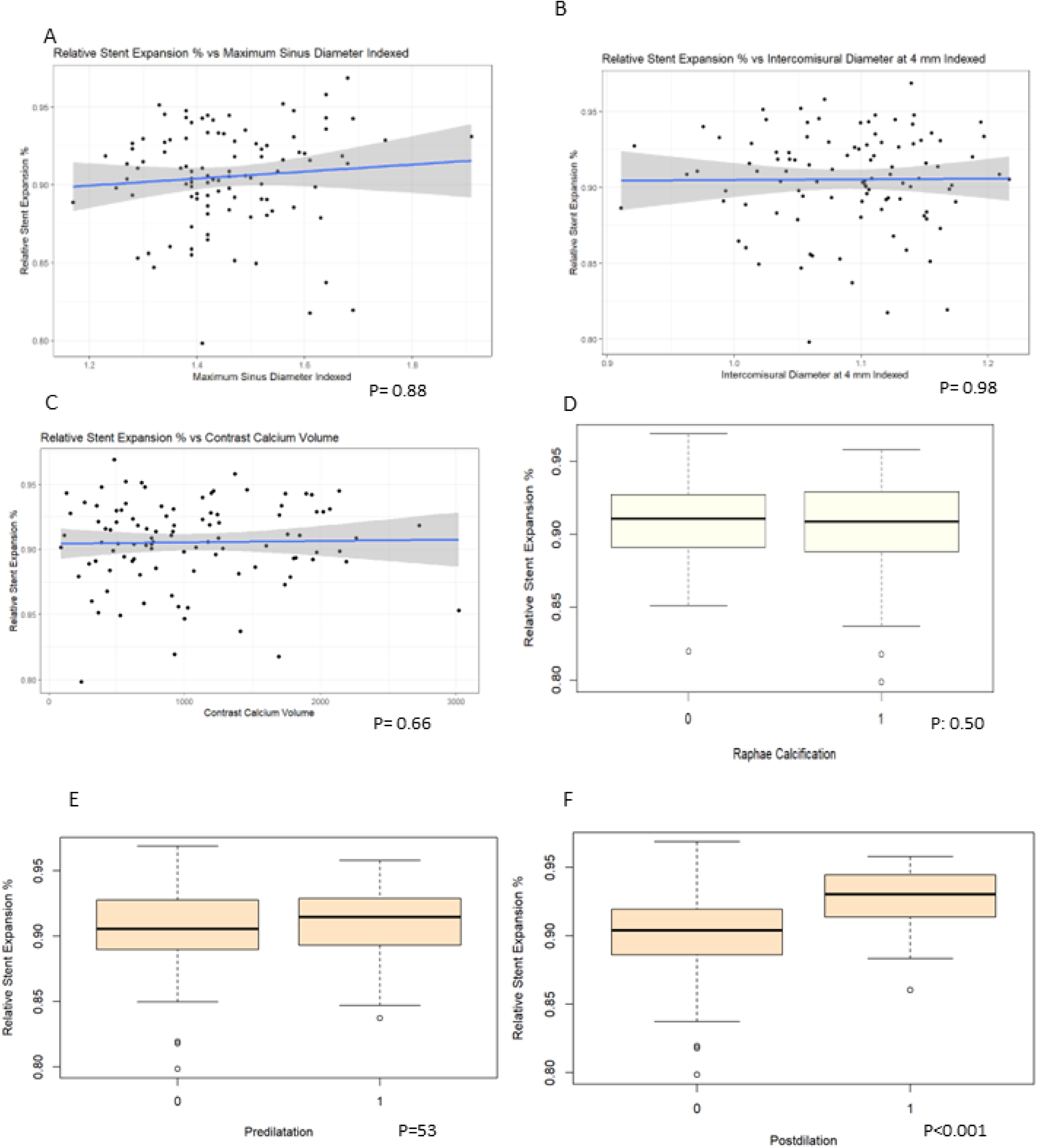
Relationship between relative stent expansion and pre-TAVR CT and procedural variables. Scatter plots for relative stent expansion vs total calcium, maximum and minimum sinus diameters indexed to the area derived annular diameter, and box/whisker plots comparing relative stent expansion in those with and without raphe calcium and with or without post dilation

**Figure 4.**
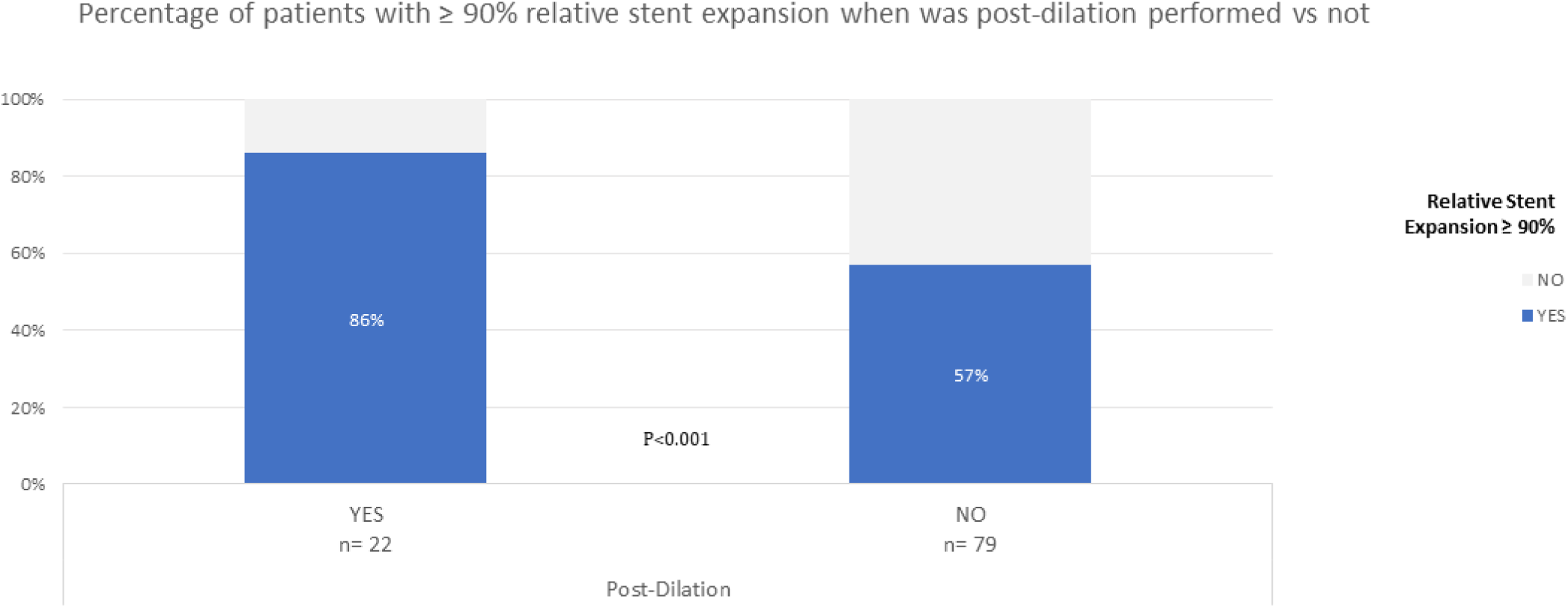
Percentage of patients ≥ 90% relative stent expansion when was post-dilation performed vs not. Association of post-dilation with the proportion of patients with ≥90% relative stent expansion.

**Figure 5:**
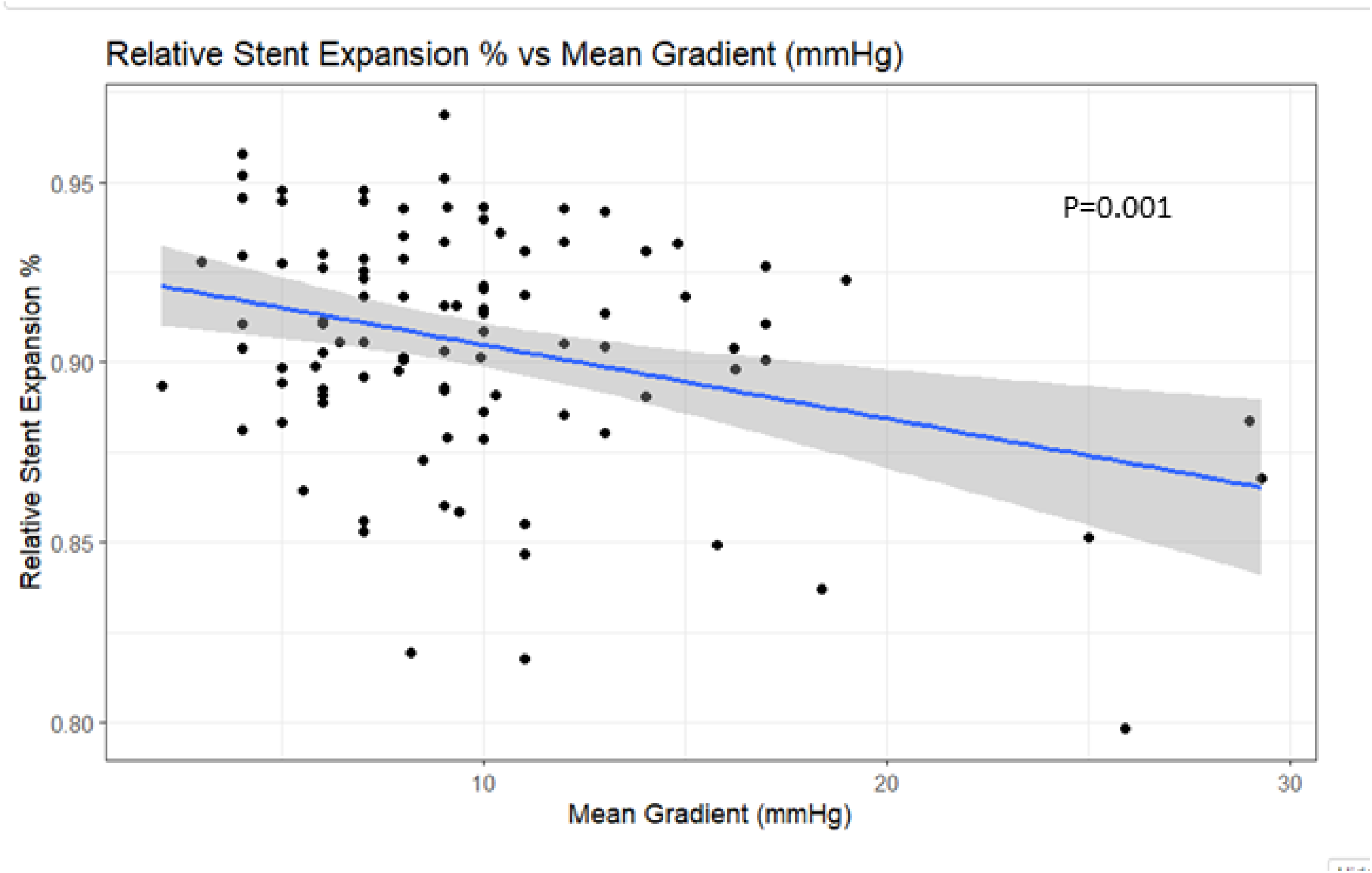
Relationship between stent expansion and post-operative hemodynamics. There was significant relationship between expansion and better post operative mean gradients with a weak negative correlation.

**Table 3:** Multivariate Analysis of Predictors of Stent Geometry.

|  | Odds Ratio | 95% Confidence Interval | P value |
| --- | --- | --- | --- |
| Relative Stent Expansion ≥ 90% | | | |
| Leaflet Calcium Volume | 0.65 | 0.39-1.05 | 0.09 |
| Presence of Raphe Calcium | 1.49 | 0.57-4.00 | 0.65 |
| Maximum Sinus Diameter indexed to annular area | 4.13 | 1.04-209.26 | 0.45 |
| Minimum Intercommisural diameter at 4 mm indexed to annular area | 31.66 | 1.36-95.21 | 0.38 |
| Performance of post-dilation | **5.55** | **1.65-25.77** | **<0.011** |
| Performance of pre-dilation | 1.52 | 0.58-4.16 | 0.39 |
| Stent Ellipticity |  |  |  |
| Leaflet Calcium Volume | 1.45 | -1.19- 1.48 | 0.82 |
| Presence of Raphe Calcium | **1.83** | **2.20-3.58** | **0.02** |
| Maximum Sinus Siameter indexed to annular area | 1.94 | -9.84- 3.10 | 0.30 |
| Minimum Intercommisural diameter at 4 mm indexed to annular area | **2.77** | **0.61 - 3.35** | **0.005** |
| Performance of post-dilation | 6.79 | -2.56 - 1.15 | 0.45 |
| Performance of pre-dilation | 7.68 | -1.46 - 1.96 | 0.76 |

#### Predictors of Stent Ellipticity

Maximum stent ellipticity ≥ 1.1 was observed in 47/101 patients and the mean stent ellipticity was 1.10 +/- 0.04 The significant predictors of stent ellipticity were the presence of raphe calcification and the ICD at 4 mm indexed to the annular area (Table 3). In contrast, there was no observable relationship between maximum sinus dimensions, leaflet calcium volume, or the performance of pre and post dilation (Figure 6). There was no relationship between stent ellipticity and post-operative gradients

**Figure 6:**
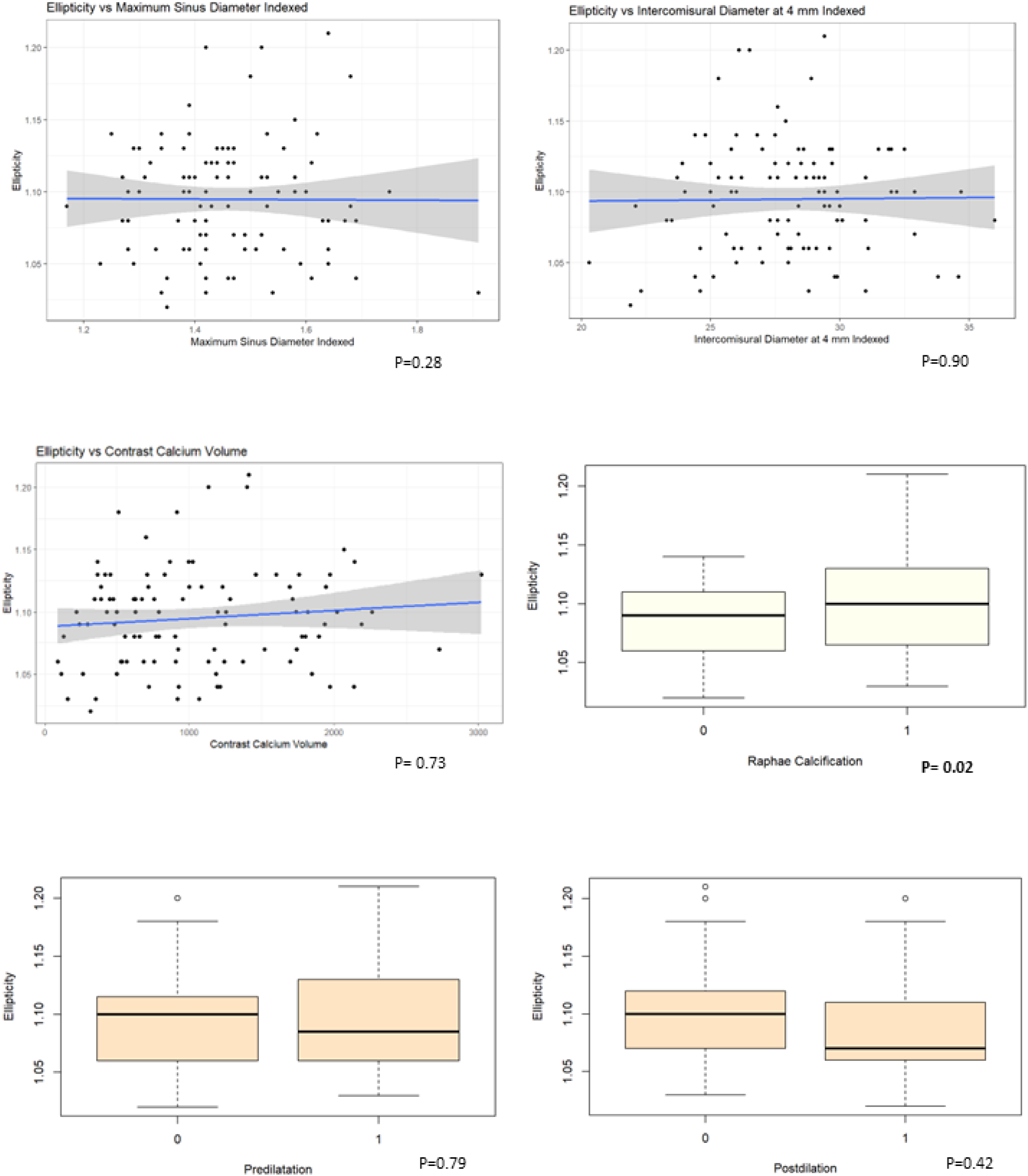
Relationship between stent ellipticity and pre-TAVR CT and procedural variables. Scatter plots for stent ellipticity vs total calcium, maximum and minimum sinus diameters indexed to the area derived annular diameter, and box/whisker plots comparing stent ellipticity in those with and without raphe calcium and with or without post dilation

Figure 7 illustrates examples of stent geometry and the associated pre-TAVR CT imaging findings. There were no difference in terms of means about expansion and ellipticity between Sievers Type 0 and Type 1 Bicuspid (p=0.658 and p=0.710 respectively).

**Figure 7:**
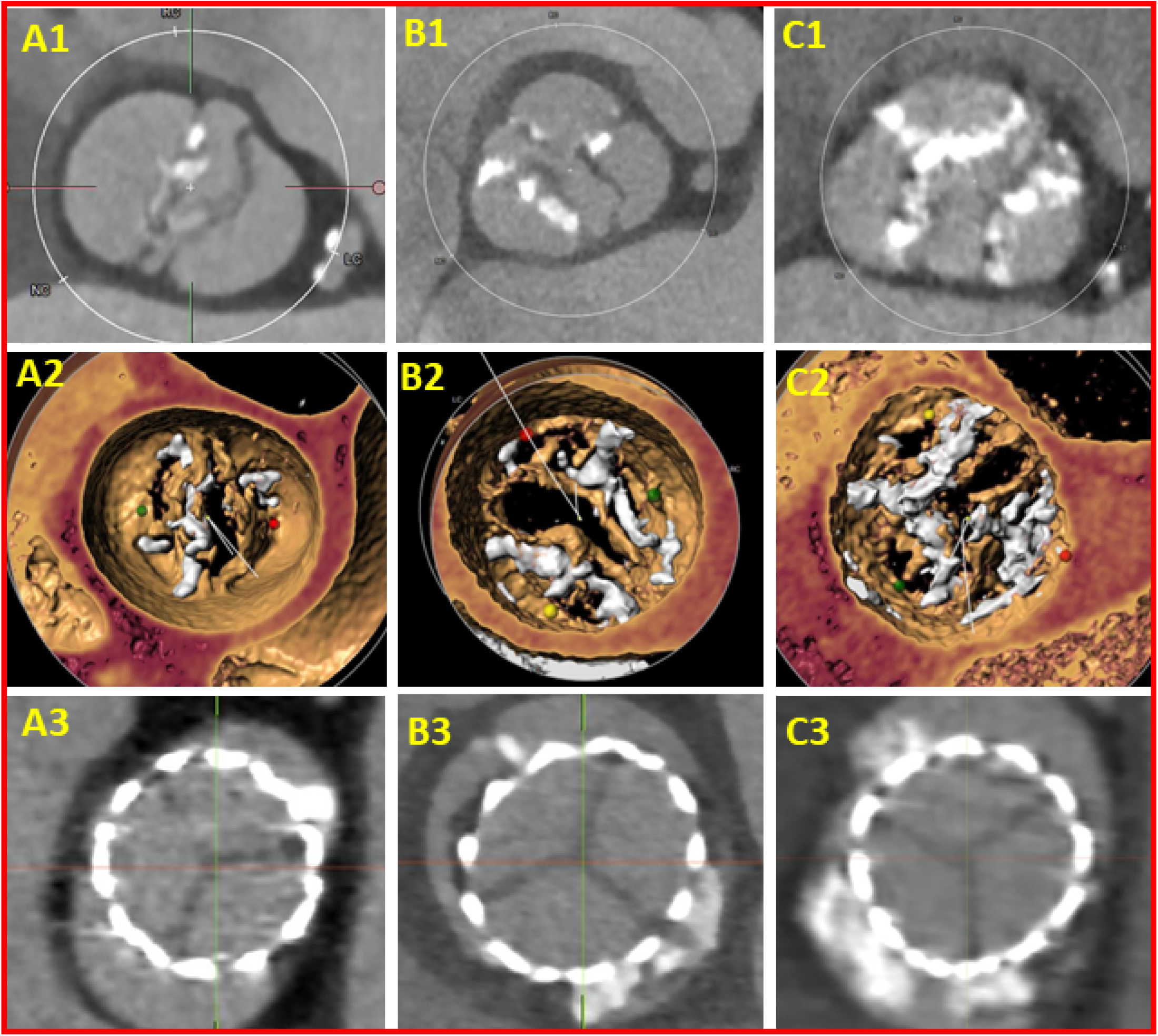
Examples illustrating stent expansion and ellipticity in patients with bicuspid valves treated with BEV. Column A, 65 years old male, with an annulus of area 516 mm2, calcium volume 500mm3, who underwent a TAVR procedure with Sapien 3 valve 26 mm + 2 cc. A3: Post stent CT shows relative stent expansion of 92.1% and ellipticity of 1.05. Column B, 75 years old female, with an annulus area of 426 mm2 and calcium volume of 513 mm3, who underwent a TAVR procedure with Sapien ultra valve 26 nominal dilation. B3: Post stent images show relative stent expansion of 90.1% and ellipticity of 1.09. Column C, 79 years old male, aortic annulus area of 664 mm2 and calcium volume of 1974 mm3, who underwent a TAVR procedure with Sapien 3 valve 29 + 2 cc. C3: Post stent Computed tomography shows relative stent expansion of 92.5% and ellipticity of 1.04.

Hypoattenuation leaflet thickening (HALT) was noted on follow-up CT in 8 out of 101 (8%). Among the 8 cases, 1 patient developed an elevation of gradient of more than 50% of the baseline gradient, however, the mean gradient remained less than 20 mmHg. There was no relationship observed between stent expansion ≥ 90% or Ellipticity ≥ 1.1 and the presence of HALT (p=0.91 and p=0.21 respectively).

## Discussion

We assessed stent geometry on post TAVR CT images in patients undergoing BEV for treatment of bicuspid valve stenosis and demonstrated (1) that the maximum distortion of stent geometry occurs near the midframe of the stent and (2) that relative stent expansion was associated with performance of post dilation while maximum stent ellipticity was associated with raphe calcification and ICD at 4 mm indexed to the annular area. Maximum Sinus dimension and valve calcium volume did not impact stent geometry.

Prior studies of BEV stent geometry on post TAVR CT have consistently shown that the minimal stent area is typically obtained at or very near to the midframe level. In a large series of tricuspid valves, the CT derived mean stent deformation index was 1.09 in patients treated with BEV^5^. This measure is the mathematical inverse of relative stent expansion used in our study and equates to mean relative stent expansion of 92%. In that study, stent eccentricity was also common and both expansion and deformation were associated with hypoattenuation leaflet thickening. Specifically in bicuspid valves, prior studies have shown relative stent expansion of 89% for bicuspid vs 91% for tricuspid valves^7^ and stent ellipticity of 1.07 to 1.14^2,7,8^. Our observation of mean relative stent expansion of 90.53% and maximum stent ellipticity of 1.10 were similar to these prior reports. Like prior studies, we observed the maximum distortion of stent geometry at or near the midframe of the stent (Figure 2), presumably due to the interaction of the stent with the native leaflets at that level.

Given that patients with bicuspid aortic stenosis are generally at low risk for surgical intervention as in our cohort (mean STS score 2.1), the identification of pre-TAVR CT variables associated with favourable or unfavourable TAVR stent geometry may aid in selection of patients for TAVR versus SAVR^11,12^. A prior registry including patients treated from 2012 to 2019 showed that above median leaflet calcification and extensive raphe calcification were associated with a substantially increased risk of peri-procedural complications^13^ However, it is likely that awareness of these risks has been incorporated into clinical practice over time and contemporary registries of TAVR treated bicuspid patients have very low complications that are similar to tricuspid pathologies^4,14^

If pre-TAVR CT findings could predict stent results, it might further refine current TAVR selection criteria, for example, by avoiding TAVR in a patient that might be highly likely to have an under-expanded stent. However, in our study, neither calcium burden nor sinus dimensions were strong predictive of post TAVR stent expansion. Post-dilation did impact relative stent expansion and was associated with a significant reduction in the proportion of patients with ≤90% relative stent expansion. Re-inflation at the same deployment volume, even in the absence of para-valvular leak, has been proposed as a method to optimize BEV deployment. These results of this study support further evaluation of this strategy. The presence of raphe calcification and ICD at 4 mm impacted on stent ellipticity. When the raphe is calcified, it likely presents a relatively fixed obstacle to stent expansion resulting in valve expansion in the contra-lateral direction and such expansion may be constrained in patients with small sinus dimensions.

## Limitations

Only patients with successful TAVR were included, therefore, there was selection bias of excluding patients who might had major complications after their TAVR procedure, although effort was made to include consecutive bicuspid patients treated with BEV, follow-up CT was performed at operator discretion and not per protocol. Therefore, non-random factors may have influenced identification of patients for follow-up imaging. Importantly, any patients undergoing CT post TAVR to investigate newly elevated gradients were excluded. Performance of post dilation was at the discretion of the operator and the comparison between those with and without post-dilation is not randomized.

## Conclusion

Among patients undergoing BEV for treatment of bicuspid valve stenosis, relative stent expansion was associated with performance of post dilation while stent ellipticity was associated with the raphe calcification and the intercommisural distance relative to annular area.

### What is known?

- Bicuspid aortic valves (BAV) with severe stenosis are generally at low risk for surgical intervention. The unique anatomic substrate of bicuspid valves has raised concern about potential suboptimal stent geometry and its impact on valve hemodynamics, durability, and repeatability.
- The identification of pre-TAVR CT variables associated with favourable or unfavourable TAVR stent geometry may aid in selection of patients for TAVR versus SAVR.

### What the study adds?

- Relative stent expansion ≥ 90% was associated with the performance of post-dilation. Stent ellipticity was associated with raphe calcification and Intercommisural distance. Maximum sinus dimensions and valve calcium volume did not impact stent expansion.
- Studies to evaluate determinants of stent geometry and the long term implications of these findings and interventional studies to test strategies to optimize valve deployment.

## Data Availability

The dicom data, and statistics outputs supporting the conclusions of this article will be made available by the authors, without undue reservation.

## Abbreviation List

TAVR: transcatheter aortic valve replacement
SAVR: Surgical aortic valve replacement
PVL: paravalvular leak
CT: computed tomography
BEV: balloon expandable transcatheter aortic valve
BSA: body surface area
BAV: Bicuspid Aortic Valve
HALT: hypoattenuation leaflet thickening

## Notes

### Competing Interest Statement

The authors have declared no competing interest.

### Funding Statement

No external funding was received for this manuscript

### Author Declarations

HiREB at HHS approved and every center locally got the respective HiREB

